# Stage-aware Brain Graph Learning for Alzheimer’s Disease

**DOI:** 10.1101/2024.04.14.24305804

**Authors:** Ciyuan Peng, Mujie Liu, Chenxuan Meng, Sha Xue, Kathleen Keogh, Feng Xia

## Abstract

Current machine learning-based Alzheimer’s disease (AD) diagnosis methods fail to explore the distinctive brain patterns across different AD stages, lacking the ability to trace the trajectory of AD progression. This limitation can lead to an oversight of the pathological mechanisms of AD and suboptimal performance in AD diagnosis. To overcome this challenge, this paper proposes a novel stage-aware brain graph learning model. Particularly, we analyze the different brain patterns of each AD stage in terms of stage-specific brain graphs. We design a Stage Feature-enhanced Graph Contrastive Learning method, named SF-GCL, utilizing specific features within each AD stage to perform graph augmentation, thereby effectively capturing differences between stages. Significantly, this study unveils the specific brain patterns corresponding to each AD stage, showing great potential in tracing the trajectory of brain degeneration. Experimental results on a real-world dataset demonstrate the superiority of our model.

## I. Introduction

Alzheimer’s disease (AD) stands out as one of the most severe neurodegenerative disorders, leading to dementia in the aging population [1]. According to the statistical data released by the World Health Organization (WHO), the number of worldwide AD patients has reached 55 million. To ensure effective and timely treatment, research on AD diagnosis and prediction is crucial. Notably, AD is a staged disease, encompassing several intermediary stages between the healthy state (e.g., normal control) and AD [2], [3]. For example, mild cognitive impairment (MCI) is one of the transitional stage of AD. It is worth noting that brain characteristics, especially the features of brain functions, would be distinct at different stages of the disease [4]. Therefore, studies focusing on AD diagnosis and prediction emphasize stage-specific analysis, such as early AD diagnosis [5].

Recently, various Artificial Intelligence (AI) and machine learning techniques (e.g., graph learning [6], [7]) have shown their effectiveness in AD detection [8]–[11]. However, most current methods fail to explore the distinctive brain patterns across different AD stages, therefore lacking the ability to trace the trajectory of disease progression. Particularly, the failure to capture stage-specific features and differences between stages can result in a decrease in model performance for both disease diagnosis and prediction tasks.

To address this issue, this paper proposes a novel stage-aware brain graph learning model, exploring the differences across AD stages. In this study, we initially construct a Brain Graph (BG) for each subject based on the information extracted from neuroimage data, such as functional magnetic resonance imaging (fMRI) data and diffusion tensor imaging (DTI) data. In a BG, each node represents a brain region of interest (ROI), and each edge indicates the connectivity between two ROIs. Then, we present a Stage Feature-enhanced Graph Contrastive Learning (SF-GCL) method, distinguishing the BGs of subjects at different AD stages. Particularly, in SF-GCL, we design a stage-based graph augmentation approach, which first extracts the specific features within each AD stage and then utilizes these features to enhance the corresponding BGs, revealing the differences between AD stages.

Significantly, this paper provides insight into the pathological mechanisms of AD, exploring the specific brain patterns of each AD stage. Moreover, our model has the potential to predict the trajectory of brain degeneration, contributing to the formulation of effective therapeutic strategies aimed at mitigating the progression of AD. The main contributions of this paper are as follows:

- This paper proposes a new stage-aware brain graph learning model, which reveals the specific brain patterns across different AD stages, effectively tracing the trajectory of AD progression.
- We design a Stage Feature-enhanced Graph Contrastive Learning (SF-GCL) method, leveraging specific features within each AD stage for graph augmentation and capturing differences between stages.
- Experiments conducted on a real-world dataset demonstrate the superiority of the proposed model.

## II. The Proposed SF-GCL

### A. Problem Definition

This paper considers the problem of stage-aware brain graph analysis. Given a subject dataset 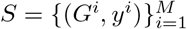, where *G*^*I*^ indicates the BG of *i*-th subject and *y*^*i*^ ∈ *Y* is the corresponding disease label, *M* denotes the total number of subjects. Notably, each BG is constructed based on the connectivities of Regions Of Interest (ROIs). Each BG is a weighted graph, denoted as *G*^*i*^ = (*V, E*^*i*^, **A**^*i*^), here 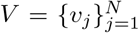 is the node set of size *N, E*^*i*^ incidates the edge set of *G*^*i*^, and **A**^*i*^ ∈ ℝ^*N×N*^ represents the weighted adjacency matrix. Note that the node set *V* is the same across subjects. The node feature matrix of *G*^*i*^ is 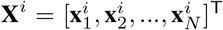, where 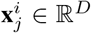 indicates the vector of node *v*_*j*_ in *G*^*i*^.

Suppose all the BGs, {*G*^1^, *G*^2^, …, *G*^*M*^ }, can be divided into *K* groups, each of which represents one stage of AD, the goal of this paper is to extract shared features within each stage, called stage features. Formally, for stage *k*, extracting the stage features **F**_*k*_ from all the group members 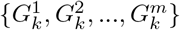, where *m* is the number of BGs in stage *k*. Then, the stage features are utilized for graph augmentation, embedding BGs into stage-specific spaces.

### B. Stage Feature-enhanced Graph Contrastive Learning

In this section, we introduce the proposed SF-GCL method (shown in Fig. 1) in detail. Particularly, SF-GCL contains two main modules: stage-based graph augmentation and contrastive objective strategy.

**Fig. 1.**
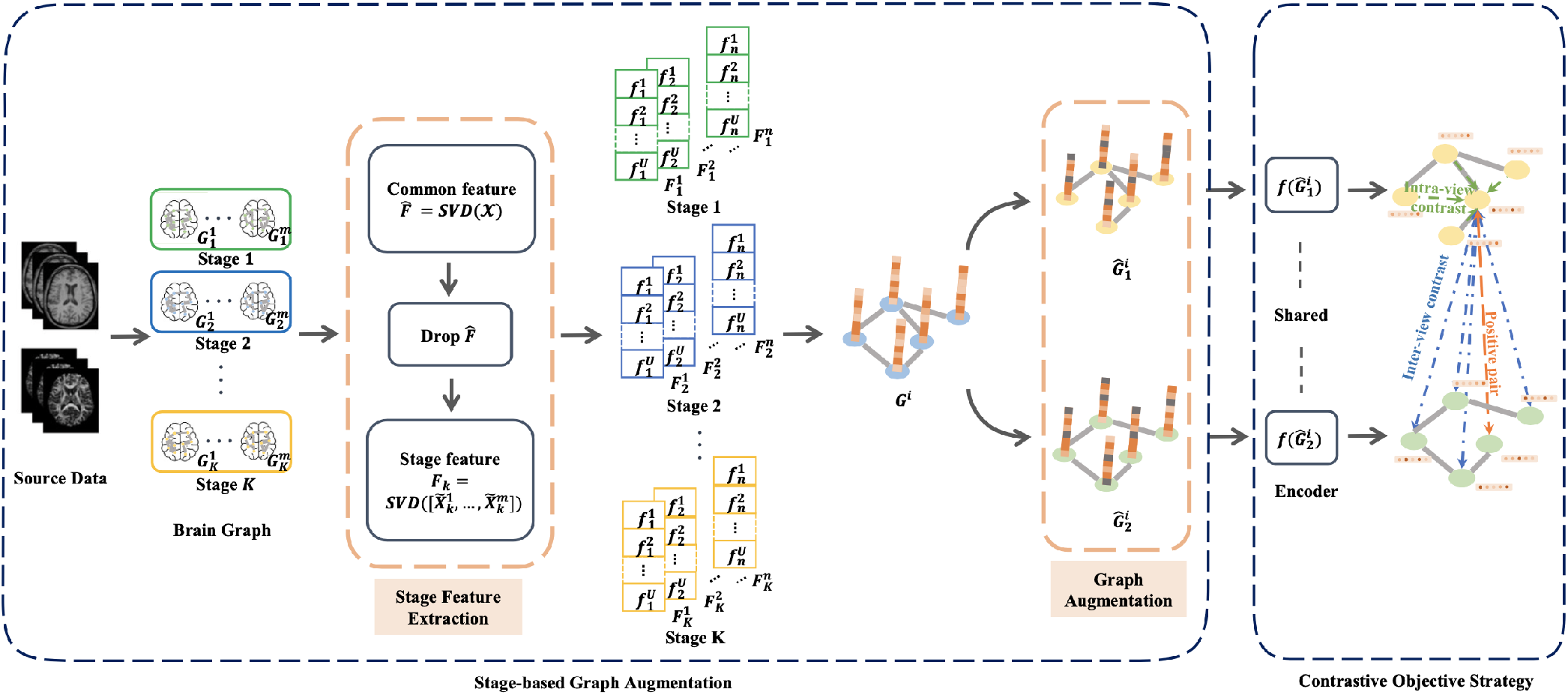
The overview of SF-GCL.

#### 1) Stage-based Graph Augmentation

A classic graph contrastive learning (GCL) model randomly generates two views of the input graph for the subsequent training process [12], [13]. Compared to other GCL models, SF-GCL employs a unique stage-based graph augmentation approach to obtain two stage-enhanced views of an input graph. Specifically, we first extract the stage features of each AD stage, then we utilize the extracted stage features to enhance the representations of corresponding input graphs, obtaining two stage-enhanced views of each graph.

##### Stage Feature Extraction

This module is designed to acquire the stage features of each AD stage. We assume that BG features exhibit variabilities across different AD stages, contributing to the differentiation of disease stages. Particularly, to prevent noise interference, we first eliminate the common features (such as features remaining the same across all the stages) of all the subjects to amplify the distinctions between stages. Afterward, the stage features 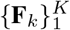 are captured using the singular value decomposition (SVD) [14], which is known for its effectiveness in dimensionality reduction and essential feature extraction based on selected top singular values.

Given a set of BGs {*G*^1^, *G*^2^, …, *G*^*M*^ }, the common features shared by all the subjects are defined as 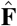. We determine whether the attributes on each dimension are shared among all subjects by assessing their commonality. The attributes with high commonality are used to form the common features. To evaluate the commonality of each attribute and learn the common features of all subjects, we take advantage of SVD:

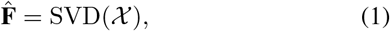

where *χ* = {**X**^1^, **X**^2^, …, **X**^*M*^ } is the node features of all the *M* subjects. SVD(·) stands as the SVD operation. We sort out node features and perform the SVD operation separately for each node to assess the commonality of their attributes. Then, we obtain common features 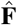 by selecting the attributes on both high and low-ranked dimensions of left singular matrices, indicating they have high commonality. Afterwards, we eliminate the common features existing in the node features of all the BGs by removing these attributes from the original node feature vectors. Thereby, we can obtain a modified node representation for each BG, denoted as 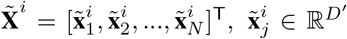, where *D′* is the modified dimension of node feature vectors.

Furthermore, we also perform SVD operation with the modified node representations of BGs at different disease stages. For stage *k* with *m* subjects, the modified node representations of BGs are denoted as 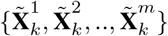. Our goal is to extract the stage-specific features of stage *k*, formulated as:

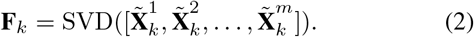

After *K* iterations, we can obtain the stage features 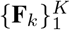, each of which corresponds to a specific AD stage. These are then used to improve the effectiveness of graph augmentation.

##### Graph Augmentation

The goal of graph augmentation is to generate two graph views by modifying the structural information without affecting the semantic labels of the input graphs [15]. Unlike many existing methods that rely on uniform data augmentation schemes, such as randomly node dropping, edge perturbation, and subgraph sampling, we introduce the stage feature-based strategy into our graph augmentation schemes. Inspired by Zhu et al. [16], who propose GCA (graph contrastive learning with adaptive augmentation) to adaptively augment the edge and node attributes of input graphs, this work adopts a stage-based graph augmentation module. Our feature masking strategy masks a fraction of less crucial attributes in node features. In particular, we assume that the stage features stand as important attributes, which should be retained.

To measure important attributes, for node *v*_*j*_, a stage weight vector **w**_*j*_ = **x**_*j*_ ⊙ **F**_*k,j*_, which is accessed by stage features, is assigned to each attribute of the node feature vector **x**_*j*_. Here, ⊙ is the element-wise multiplication, **F**_*k,j*_ indicates the stage feature vector of node *v*_*j*_. Essentially, attributes with higher stage weight values are deemed more informative and crucial. Consequently, during the augmentation process, these attributes will have a lower probability of being masked. In order to unify the value of probability, we apply a normalization based on the stage weights. For *u*-th dimension of node *v*_*j*_, we calculate the probability as:

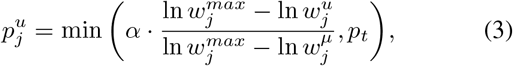

where *α* is a hyperparameter to adjust the magnitude of feature augmentation, 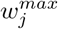 and 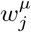 are the maximum and the average weight values of all dimensions in node *v*_*j*_, and *p*_*t*_ is the limitation of the highest probability.

Afterwards, for node *v*_*j*_, the attribute augmentation strategy leverages a random vector **b**_*j*_ ∈ {0, 1}^*D′*^ where each dimension is described by a Bernoulli distribution, such as 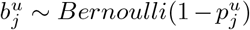. Thereby, for *G*^*i*^, the generated node features are computed by:

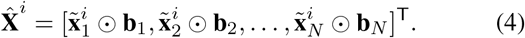

Finally, two augmented graph views 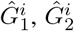 can be generated by performing our stage-based graph augmentation strategy.

#### 2) Contrastive Objective Strategy

The contrastive objective is formulated to maximize the mutual information between node embeddings and a global summary embedding [17]. Specifically, the purpose of the strategy is to distinguish the embeddings of the same node in two different views from those of other node embeddings. The node features are fed into a graph neural network-based view encoder network *f* (·) to generate embeddings containing both structure and attribute information in the views, denoted as **Z** ∼ *f* (*Ĝ*^*i*^). For any node, we designate its embedding **z**_*i,n*_ as an anchor point in one view, and its embedding in another view as a positive sample. Naturally, embeddings of other nodes in these two views are treated as negative samples. We adopt the InfoNCE [18] loss as the contrastive loss function, defined as:

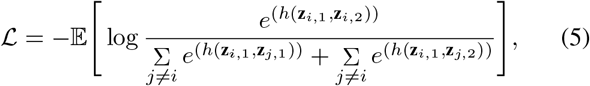

where *h*(·) is a score function that measures the similarity between two representations. In the denominator, the left part represents the sum of intra-view negative pairs, while the right part signifies the sum of inter-view negative pairs. The theoretical optimization objective is to minimize the value of the contrastive loss *ℒ*, which is equivalent to maximizing a lower bound on the mutual information between positive pairs of views.

## III. Experiments

### A. Datasets

The data utilized in this study is sourced from the AD Neuroimaging Initiative (ADNI)^1^ database, including various neuroimaging data (e.g., DTI and fMRI). Based on the collected DTI and fMRI data from ADNI, we construct BGs by computing the Pearson Correlation Coefficient between ROIs.

The collected dataset comprises a total of 460 subjects, categorized as normal control (NC) (*m* = 211), MCI (*m* = 195), and AD (*m* = 54), carefully matched for both age and sex ratio. The subjects, ranging from 49 to 96 years old, willingly undergo all testing procedures, and participate in longitudinal surveys. Table I summarizes the number of subjects in each group along with their corresponding scores on the minimental state examination (MMSE) and clinical dementia rating (CDR), which are used to assess cognitive function and the severity of AD. Scores of MMSE range from 0 to 30, with higher scores indicating better cognitive function. The CDR is typically employed to evaluate the severity of dementia, where 0 signifies normal, 1 denotes mild dementia, 2 indicates moderate dementia, and 3 represents severe dementia. Particularly, we conduct experiments for graph classfication task based on the DTI data of all 460 subjects, and the fMRI data of 195 MCI and 54 AD subjects is collected for graph clustering task.

**TABLE I.**
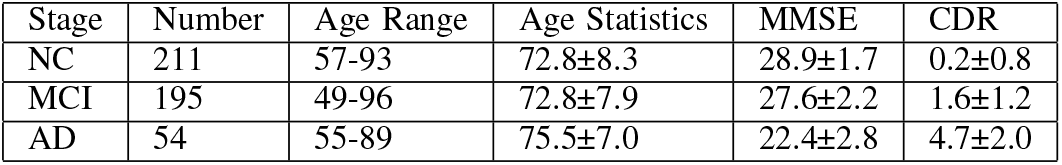
Clinical and demographic information of subjects in ADNI dataset.

| Stage | Number | Age Range | Age Statistics | MMSE | CDR |
| --- | --- | --- | --- | --- | --- |
| NC | 211 | 57-93 | 72.8±8.3 | 28.9±1.7 | 0.2±0.8 |
| MCI | 195 | 49-96 | 72.8±7.9 | 27.6±2.2 | 1.6±1.2 |
| AD | 54 | 55-89 | 75.5±7.0 | 22.4±2.8 | 4.7±2.0 |

### B. Results

To assess the effectiveness of SF-GCL, we conduct graph classification and graph clustering tasks based on DTI and fMRI data, respectively. Fig. 2 shows the accuracy and F1 score of graph classification task on DTI data. Comparing with GCA model, which ignores AD stage features, the accuracy of SF-GCL improves about 3.43%, achieving 60.00 *±* 2.48%. In addition, the F1 score of SF-GCL is about 2.80% higher than GCA, with 58.33 *±* 1.52%. For graph clustering task, we mainly distinguish two different AD stages (MCI and AD) based on fMRI data. Fig 3 visualizes the graph clustering result, showing that our model accurately maps BGs to stage-specific spaces.

**Fig. 2.**
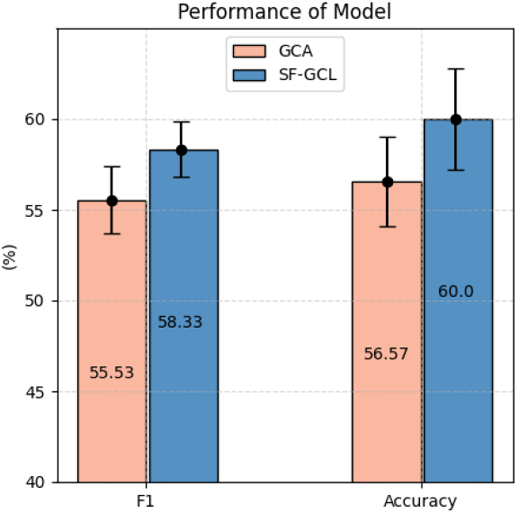
The accuracy and F1 score of graph classification task on DTI data.

**Fig. 3.**
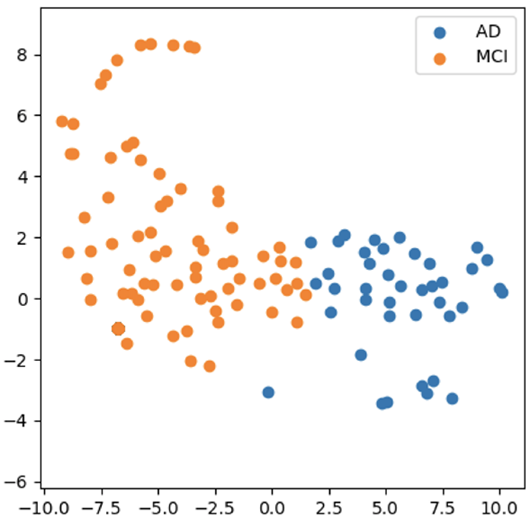
Graph clustering result on fMRI data.

Overall, the experimental results demonstrate that our model excels in capturing the non-linear relationship between input features and target variables. In both graph classification and graph clustering tasks, SF-GCL shows its superiority in distingushing different AD stages. These findings further affirm the outstanding performance of our model in highlighting the specific brain patterns across AD stages, providing robust support for tracing the trajectory of AD progression.

## IV. Conclusion

This paper proposes a new stage-aware brain graph learning model to anaylze different brain graphs across AD stages. Particularly, we present a Stage Feature-enhanced Graph Contrastive Learning method (SF-GCL), where AD stage features are extracted to perform graph augmentation, thereby effectively capturing differences between AD stages. Then, we conduct experiments on a real-world dataset for graph clustering and graph classification tasks. Experimental results show the superiority of our model, and highlight its potential in tracing the trajectory of brain degeneration. Future work could expand to a more comprehensive classification of AD stages, such as further subdividing MCI into early-stage and late-stage categories to facilitate a more precise tracking of the disease.

## Data Availability

All data produced in the present study are available upon reasonable request to the authors

## Footnotes

1 https://adni.loni.usc.edu/

## Notes

### Competing Interest Statement

The authors have declared no competing interest.

### Funding Statement

This study did not receive any funding

### Author Declarations

The study used ONLY openly available human data that were originally located at the AD Neuroimaging Initiative database

